# The first month of the COVID-19 outbreak in 46 sub-Saharan African countries. A comparative analysis of growth rates

**DOI:** 10.1101/2020.04.09.20057091

**Authors:** Martin J. Prince, Atalay Alem, Dixon Chibanda, Lara Fairall, Abebaw Fekadu, Charlotte Hanlon, Crick Lund, Andy Leather, Inge Petersen, Ruth Verhey, Haja Wurie

## Abstract

**Background:** The COVID-19 outbreak in sub-Saharan African countries started after those in Asia, Europe and North America, on 28^th^ February 2020. The susceptibility to infection of populations in that region has been debated. Outbreaks on the scale of those seen elsewhere would pose substantial challenges. There are reasons for concern that transmission may be high and difficult to control, rapidly exceeding capacity to meet the needs for hospitalization and critical care.

**Methods:** We obtained data on daily new confirmed cases for all 46 countries from the World Health Organization, and used these to model and visualize growth trajectories using an AutoRegressive Integrated Moving Average (ARIMA) model. We then estimated doubling times from growth rates estimated from Poisson regression models, and by back counting from the most recent observation. We also calculated the time from 1^st^ to 50^th^ case, and the time from 5^th^ to 100^th^ case. These indicators were compared with the same summary indicators of growth at the same stage of the outbreak in highly affected European countries.

**Results:** Kenya was the only country with clear evidence of exponential growth. Nineteen countries had either reported no cases, were in the first few days of the outbreak, or had reported fewer than 10 cases over a period of two or more weeks. For the remaining 27 countries we identified four growth patterns: slow linear growth, more rapid linear growth, variable growth patterns over the course of the outbreak, and early signs of possible exponential growth. For those in the last three groups, doubling times ranged from 3 to 4 days, times from 1^st^ to 50^th^ case from 12 to 29 days, and from 5^th^ to 100^th^ case from eight to 15 days. These early indicators are comparable to those in European countries that have gone on to have substantial outbreaks, and time to 50^th^ case was shorter suggesting lesser effectiveness of contact-tracing and quarantine in the early phase.

**Conclusion:** The 46 sub-Saharan African countries, home to over one billion people, are at a tipping point with clear potential for the outbreak to follow a similar course as in HIC in the global north. Radical population-level physical distancing measures may be required, but their impact on poor, disadvantaged and vulnerable people and communities need mitigating. Health systems in the region need urgent technical and material support, with testing, personal protection, and hospital/ critical care.

## BACKGROUND

Just over one billion people (14% of the world’s population) live in the 46 countries of the Sub-Saharan African region (SSA). The first confirmed case of COVID-19 infection in the region was reported, in Nigeria, on the 28th February 2020 [1], just two months after the first notification of a pneumonia of unknown cause in Wuhan, China. On the 11th March 2020 the Director General of the World Health Organization, Dr Tedros Adhanom Ghebreyesus, classified the COVID-19 outbreak as a global pandemic. By 1st April 2020 43 of 46 sub-Saharan Africa countries had reported confirmed cases of COVID-19. The pandemic had arrived in sub-Saharan Africa.

Management of the pandemic in SSA poses unique challenges [2], although with much inter-country variation in baseline infection prevention and control preparedness and capacity, partly linked to recent experience with other widespread outbreaks, e.g Ebola. Demographics are favourable; population ageing is at an early stage with just 3% aged 65 years and over, while close to half the SSA population are aged under 20 years [3]. The theoretical benefits of the warmer climes in equatorial and tropical countries on COVID-19 transmission are yet to be convincingly demonstrated [3]. However, SSA as a region, has the world’s greatest concentration of least developed countries [4], fragile and conflict-affected states [5], morbidity [6], inadequate housing [7], and weak healthcare systems [8]. Those living in poverty are likely to be more susceptible to COVID-19 infection and its effects, due to overcrowding, poor sanitation, food insecurity and undernutrition, low education and health literacy, and a high underlying disease burden including from HIV and tuberculosis. To add to the perfect storm, hospital beds are scarce and inequitably distributed [9], and critical care is limited to some tertiary care and private facilities, with few ventilators, limited oxygen supplies, and little obvious scope for rapid expansion [10,11]. Testing capacity was limited at the start of the outbreak [1], and has not expanded sufficiently to keep pace with demand. Infection prevention and control preparedness needs further strengthening, at every level of the system. Any weaknesses in health sector governance and leadership, and integration/ communication from Federal, to Provincial and District levels will be exposed. If there is widespread community transmission, the case fatality rate may well exceed that in other regions.

For all these reasons, it is important to monitor the outbreak across sub-Saharan Africa, comparing patterns of transmission with those of other countries that preceded it, for clues that this may give as to the future course of the outbreak in the African region. In a recent report on the COVID-19 outbreak in West Africa [12], the authors sought to compare growth in numbers of cases over the initial phase of the outbreak with those in selected West African Countries, and Ethiopia and Egypt. Their conclusion was that in two West African Countries, Burkina Faso and Cote d’Ivoire, the early course was like that in European countries that have gone on to have very substantial outbreaks that challenged the capacity even of their advanced health systems to deliver critical care.

At the National Institute of Health Research Global Health Research Unit on Health Systems Strengthening in Sub-Saharan Africa, we have been collating and disseminating information on daily confirmed cases of COVID-19 infection, and deaths for all 46 sub-Saharan African countries. Our aim is to document and study the contextual impact of health system indicators, health system responses (including testing capacity and policies) and transmission control measures, while providing a platform for news and views from those involved and affected across the region. Accordingly, we have set out here to document the course of the outbreak across the 46 countries over the first month (36 days from February 28^th^ 2020 to April 3^rd^ 2020) since the first reported confirmed case in the region. Specifically, we have tried to explore the applicability and utility of the ‘doubling time’ indicator to compare growth trajectories across countries, and other approaches meaningfully to compare growth rates with those at similarly early stages of the outbreaks in European countries.

## METHODS AND RESULTS

Our data resource comprises data on daily new confirmed cases, deaths among confirmed cases, and cumulative cases and deaths, obtained from the World Health Organization retrospectively on the 28^th^ March 2020, and contemporaneously thereafter. We use these data to calculate a cumulative attack rate on each day (per 100,000 population) and case fatality rate (number of deaths among confirmed cases divided by number of confirmed cases, expressed as a percentage). Cleaned data with derived variables are available for download at ##### in both wide and long format (suitable for time series analysis). The focus of the analyses reported in this paper is upon the course of the outbreak in terms of cumulative confirmed cases.

Four of the 46 sub-Saharan African countries (Comoros, Lesotho, Sao Tome and Principe, and South Sudan) were excluded from this analysis as they had not reported any cases to 3^rd^ April 2020. Four further countries were excluded since they have yet to report 10 or more cases, and are in the very early stages of their outbreaks – Burundi and Sierra Leone (two cases each in three days, Malawi (three cases in two days), and Botswana (four cases in three days). There is a further group of 11 countries (Table 1) with low numbers of confirmed cases more than one to three weeks after the start of the outbreak. All show slow to negligible linear growth, and are also excluded from the analysis.

**Table 1.**
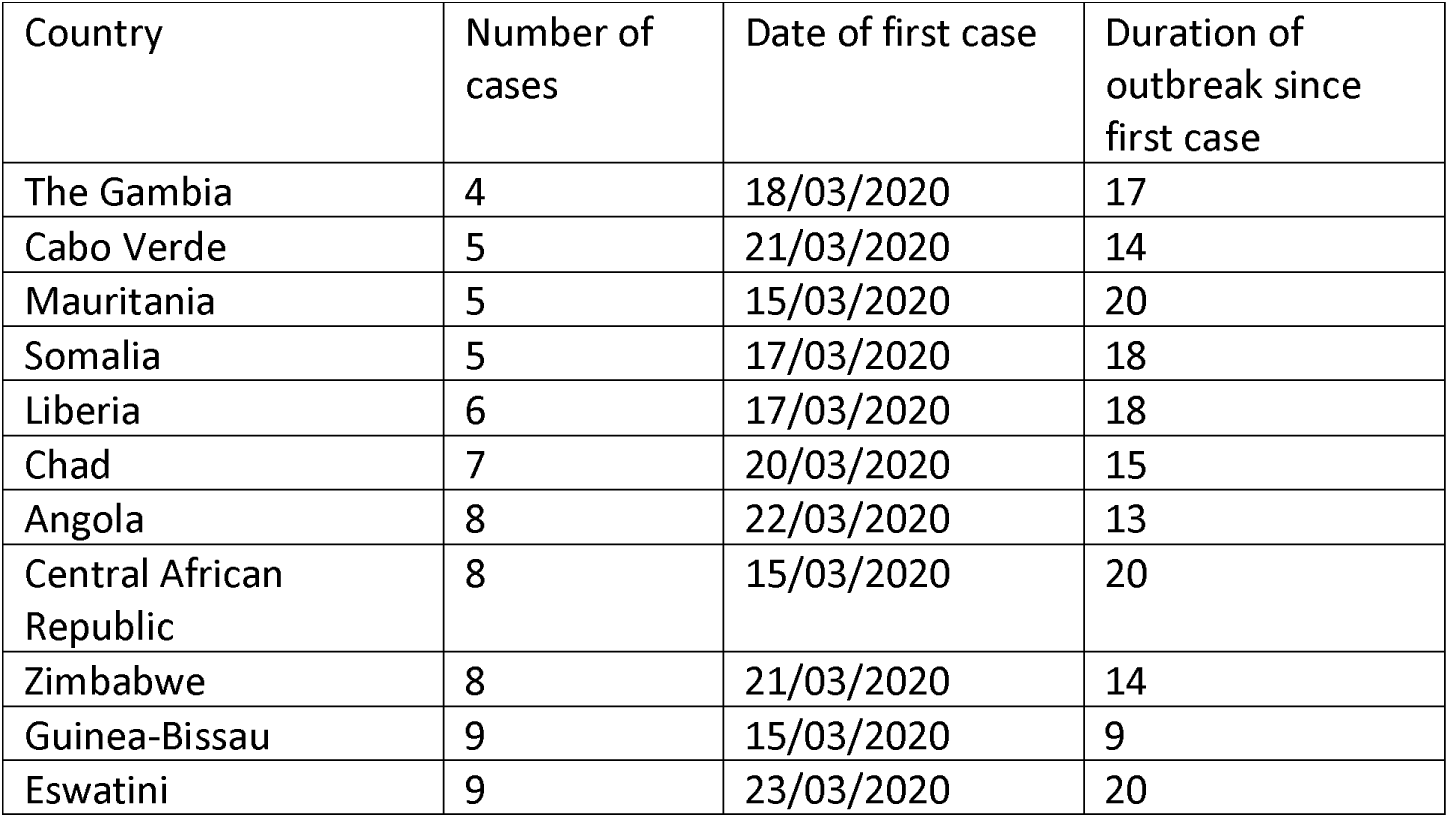
Sub-Saharan African countries with fewer than 10 reported confirmed cases of COVID-19 infection by 03/04/2020

The remaining 27 countries are a median of 20 days into the outbreak (range 9 to 36 days), and have confirmed a median of 44 cases (range 10 to 1462 cases)

For these 27 countries, we first explored the growth patterns in the time series data (daily cumulative total of confirmed cases) by fitting, separately for each country, an AutoRegressive Integrated Moving Average (ARIMA) model. ARIMA models capture a suite of different standard temporal structures in time series data. We explored the possibility of a periodic effect, hypothesising fewer cases reported at or after weekends, but found none. The integrated component establishes whether there is a trend in the lagged differences in data over time that needs to be accounted for in the model fit, and if so, whether this is ‘first order’ (a linear trend, with cases increasing by a constant number each day) or ‘second order’ (exponential growth with numbers increasing by a constant multiplier each day). The optimal ARIMA models fitted adequately for all countries. However, in only one country, Kenya, did a second order difference setting (ARIMA 0,2,0 - exponential growth) improve model fit over a first order setting (ARIMA 0,1,0 – linear growth). The concept of ‘doubling time’ assumes exponential growth, with a constant growth rate (mu) per unit of time.

In seven other countries (Burkina Faso, Cote d’Ivoire, Democratic Republic of Congo, Nigeria, Senegal and South Africa) the optimal model was natural logarithm transformed to fit to a linear growth pattern. Therefore, for these countries there may be some evidence of a tendency towards exponential growth. Closer inspections of these and other countries’ growth trajectories suggested four common patterns (see Figure 2 for examples and Table 2 for a full breakdown)

**Table 2.** Doubling time in days for increase in cumulative number of cases in sub-Saharan African countries to 03/04/2020

| Country by growth pattern | Doubling time in days (recency method) | Doubling time in days (Poisson model) | Cases to 03/04/2020 | Duration of outbreak (in days) | Days to 50<sup>th</sup> case, after 1<sup>st</sup> case | Days to 100<sup>th</sup> case, after 5<sup>th</sup> case |
| --- | --- | --- | --- | --- | --- | --- |
| Slow linear |  |  |  |  |  |  |
| Benin | 3.7 | 5.0 (3.8-7.5) | 13 | 18 | n/a | n/a |
| Ethiopia | 6.3 | 5.9 (4.9-7.3) | 31 | 21 | n/a | n/a |
| Equatorial Guinea | 4.7 | 5.9 (4.6-8.2) | 15 | 20 | n/a | n/a |
| Gabon | 1.0 | 6.9 (5.2-10.5) | 18 | 21 | n/a | n/a |
| Mozambique | 7.0 | 4.9 (3.2-9.7) | 10 | 12 | n/a | n/a |
| Namibia | 8.3 | 6.0 (4.9-8.7) | 13 | 20 | n/a | n/a |
| Tanzania | 11.3 | 6.5 (5.1-8.8) | 20 | 18 | n/a | n/a |
| Togo | 9.0 | 4.8 (4.3-5.4) | 39 | 28 | n/a | n/a |
| More rapid linear |  |  |  |  |  |  |
| Burkina Faso | 8.5 | 4.3 (4.1-4.6) | 261 | 24 | 12 | 8 |
| Cote d'Ivoire | 5.9 | 3.5 (3.3-3.7) | 190 | 23 | 14 | 12 |
| Djibouti | 4.5 | 2.1 (1.9-2.5) | 41 | 17 | n/a | n/a |
| Democratic Republic of Congo | 5.0 | 3.9 (3.6-4.2) | 134 | 24 | 16 | 14 |
| Madagascar | 4.7 | 4.2 (3.6-5.1) | 65 | 13 | 13 | n/a |
| Rwanda | 8.0 | 4.8 (4.4-5.4) | 84 | 20 | 13 | n/a |
| Senegal | 8.1 | 5.1 (4.9-5.4) | 195 | 32 | 20 | 14 |
| Uganda | 4.5 | 3.4 (2.8-4.1) | 44 | 13 | n/a | n/a |
| Variable rates of increase |  |  |  |  |  |  |
| Congo | 2.3 | 3.7 (3.2-4.5) | 41 | 20 | n/a | n/a |
| Ghana | 8.4 | 3.8 (3.5-4.0) | 204 | 21 | 12 | 9 |
| Mali | 5.4 | 3.0 (2.3-4.4) | 28 | 9 | n/a | n/a |
| South Africa | 8.5 | 4.0 (3.9-4.0) | 1462 | 29 | 11 | 9 |
| Niger | 1.5 | 1.9 (1.7-2.2) | 74 | 15 | 14 | n/a |
| Zambia | 5.8 | 3.0 (2.6-3.5) | 39 | 17 | n/a | n/a |
| Recent exponential |  |  |  |  |  |  |
| Cameroun | 3.4 | 3.9 (3.7-4.1) | 246 | 29 | 19 | 14 |
| Eritrea | 2.4 | 3.0 (2.3-4.4) | 20 | 13 | n/a | n/a |
| Guinea | 1.3 | 2.8 (2.4-3.3) | 52 | 21 | 21 | n/a |
| Niger | 1.5 | 1.9 (1.7-2.2) | 74 | 15 | 14 | n/a |
| Nigeria | 3.5 | 3.7 (3.5-3.9) | 174 | 36 | 29 | 13 |
| Zambia | 5.8 | 3.0 (2.6-3.5) | 39 | 17 | n/a | n/a |
| Good ARIMA model fit for exponential growth |  |  |  |  |  |  |
| Kenya | 2.5 | 3.5 (3.2-3.9) | 110 | 21 | 18 | 15 |

**Figure 1.**
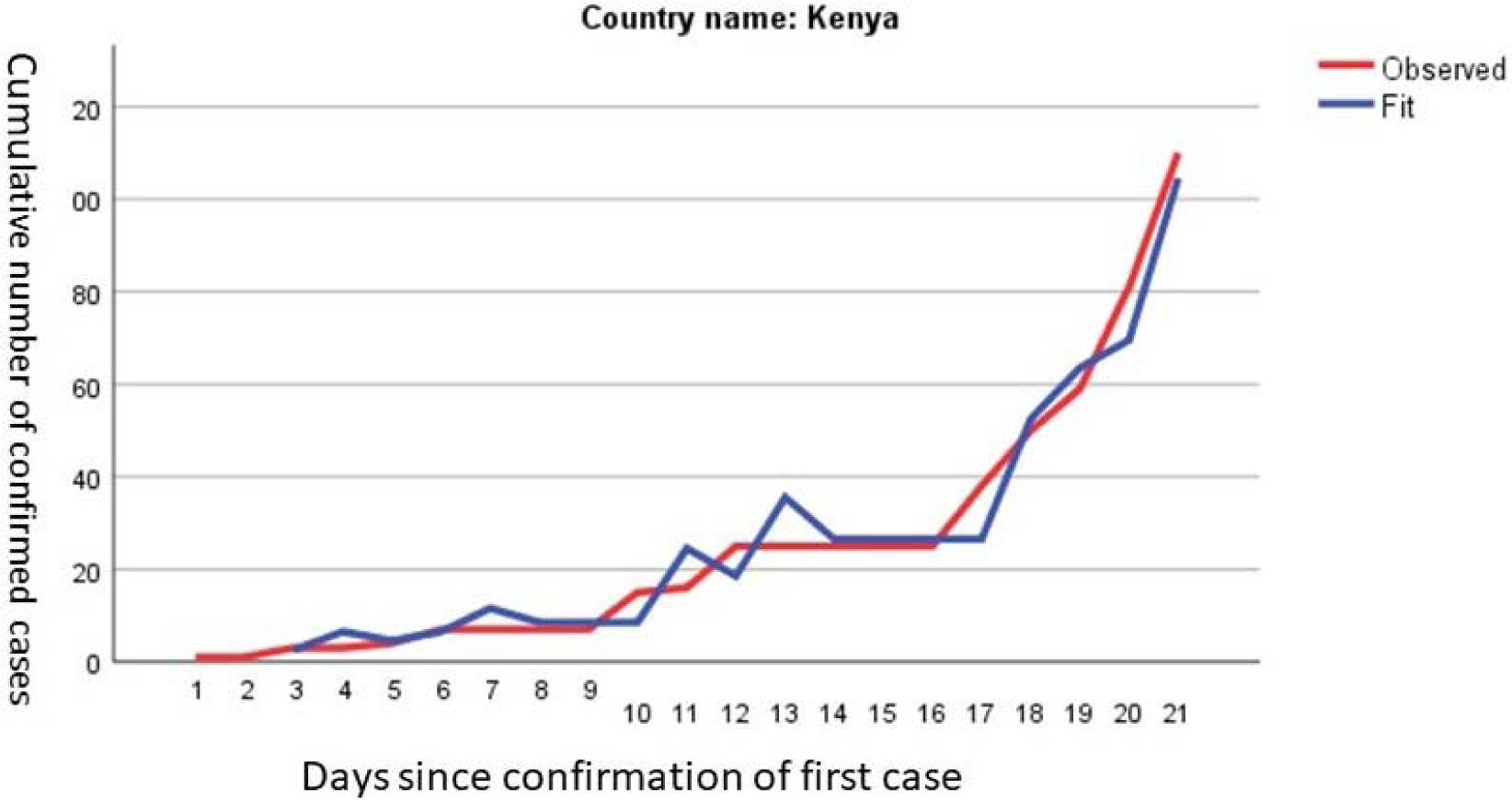
Illustration of an exponential growth trajectory, over the first 21 days of the outbreak in Kenya.

**Figure 2.**
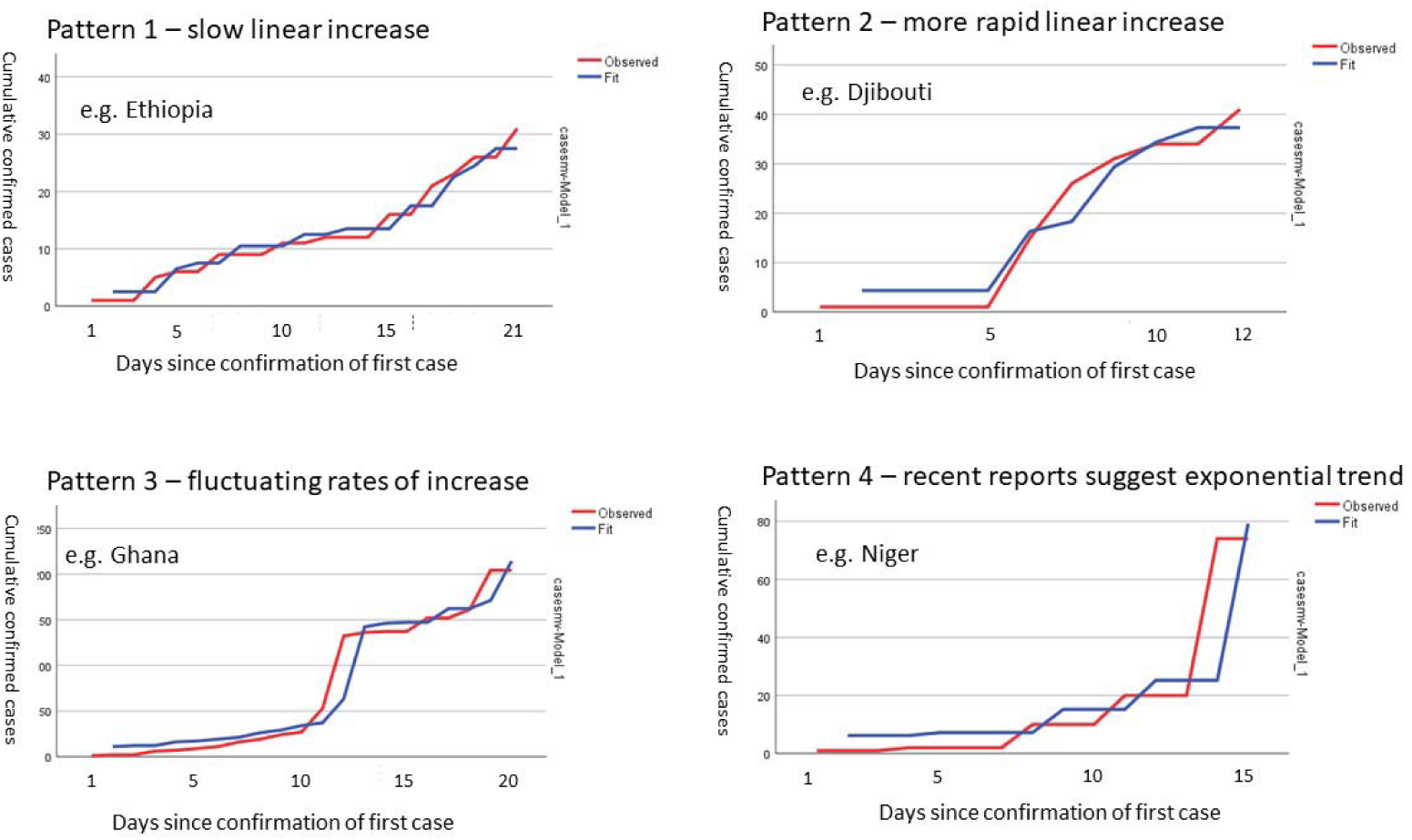
examples of different patterns of growth in cumulative confirmed cases in Sub-Saharan African countries.

Pattern 1 – a relatively slow linear increase (exemplified by Ethiopia, also noted for Benin, Equatorial Guinea, Gabon, Mozambique, Namibia, Tanzania, and Togo)

Pattern 2 – a more rapid linear increase (exemplified by Djibouti, the other examples being Burkina Faso, Cote d’Ivoire, Djibouti, Democratic Republic of Congo, Madagascar, Rwanda, Senegal and Uganda).

Pattern 3 – rates of increase in numbers wax and wane markedly over different periods, with an overall linear growth pattern. This is exemplified in Fig 2 by Ghana, the other countries fitting this pattern are Congo, Mali and South Africa.

Pattern 4 – while there is a general linear trend, the most recent reports suggest the possible beginnings of an exponential phase (exemplified by Niger, also noted for Cameroon, Eritrea, Guinea, Nigeria and Zambia).

Doubling times were estimated using two different approaches

1. from a Poisson regression, with time in days since the start of the outbreak used to estimate the daily growth rate (mu), assumed to be a constant multiplier. From this, the doubling time in days can be calculated, as 0.693/mu, with its 95% confidence intervals.
2. as proposed by the ‘Our World in Data’ group [13], the number of days taken to progress from half the number of cases current on 3^rd^ April to that number on that date. We refer to this as the ‘recency’ approach. It prioritises recent growth in numbers rather than the pattern of growth over the whole outbreak. It is not possible to calculate confidence intervals, and it is susceptible to noise from recent ebbs and flows in reporting of confirmed cases.

We also provide quantifiable indicators inspired by the graphical representations in the comparative report on the outbreak in West Africa [12]; the time in days from the first reported case to the 50^th^ case, and the time in days to the 100^th^ case after the 5^th^ case, and compare these with the same indicators derived from the COVID-19 outbreaks in the most affected European countries; Italy, France, Germany, Spain and the United Kingdom.

The stratification of countries based upon the form of the trajectory (from ARIMA models and visual inspection) was clearly related to the doubling times, more so when estimated from Poisson regression than when using the ‘recency’ approach. Doubling times were longest (reflecting slower progression) for the slow linear group (4.8 to 6.9 days), followed by the rapid linear group 2.1 to 5.1 days). Doubling times for the trajectories showing some indications of recent exponential growth, and those with variable course mainly clustered between three and four days, and that for Kenya, which showed a clear exponential trajectory, was 3.5 days (95% CI 3.2-3.9 days)

For the 13 countries to have reached >=50 cases (Burkina Faso, Cameroun, Cote d’Ivoire, Democratic Republic of Congo, Ghana, Guinea, Kenya, Madagascar, Niger, Nigeria, Rwanda, Senegal, South Africa) time from 1^st^ case to 50^th^ case ranged from 12 to 29 days. For the nine countries to have reached >=100 cases (Burkina Faso, Cameroun, Cote d’Ivoire, Democratic Republic of Congo, Ghana, Kenya, Nigeria, Senegal, and South Africa), time from 5^th^ case to 100^th^ case ranged from eight to 15 days. From data obtained from the World Health Organization COVID-19 Outbreak Situation Dashboard, the same indicators for European nations were; for time from 1^st^ case to 50^th^ case - Italy 24 days, France 35 days, Germany 31 days, Spain 32 days, United Kingdom 34 days; for time from 5^th^ case to 100^th^ case - Italy two days, France 32 days, Germany 31 days, Spain six days, and United Kingdom 24 days.

## DISCUSSION

In this report we have estimated growth rates, as doubling times, for 27 sub-Saharan African (SSA) countries using two approaches, one modeled from the entirety of the outbreak data, the other using a simpler approach based on counting back from the most recent cumulative case report to identify the date on which one half that number of cumulative cases was reported. We could not report growth rates for eight countries, which either had yet to report a case, or had only done so within the past three days. A further group of 11 countries had reported fewer than 10 cases, despite being several weeks into the outbreak. For the remaining 27 countries we identified four patterns of growth trajectory with implications for doubling times; slow linear growth (n=8, high doubling time, slow progression), more rapid linear growth (n=8, moderate doubling time, moderate progression), and variable growth (n=4), or recent growth suggestive of transition to exponential phase (n=6), or exponential growth (n=1) (low doubling time, rapid progression). For those countries that had progressed to >=50 cases, time from 1^st^ to 50^th^ case was notably shorter than in the European nations most affected to date. For those which had progressed to >100 cases, time from 5^th^ case to 100^th^ case was intermediate between the most rapidly developing outbreaks in Italy and Spain, and the outbreaks in France, Germany and the United Kingdom that, initially, grew at a slower rate.

Summarising the growth rate, and future growth potential of infectious disease epidemics in the early phase of outbreaks is important but challenging. One objective is to distinguish, as early as possible, whether and when the outbreak is transitioning from linear to exponential growth. Visual inspection, and model fitting provide useful indications. The ‘recency’ approach for estimating doubling time, widely reported on web platforms fails to address this issue, and the indicator is both crude and strongly influenced by noisy data arising from vagaries in the case detection and reporting processes. At the same time, it is important to acknowledge that modeled growth rates are assumed to be constant, a condition that is met only in the exponential growth phase, which most SSA countries have yet clearly to enter. Nevertheless, the doubling times estimated from Poisson regression, with their 95% confidence intervals, do seem to provide a useful stratification of variations in growth of confirmed cases across the region. They should not, however, be interpreted concretely.

Eleven SSA countries have shown negligible growth two to three weeks into the outbreak, and a further eight countries are showing slow linear growth over similar periods. This may or may not be reassuring. The most affected European countries also showed negligible growth in confirmed cases over most of the first month of the outbreak. In two of those countries, Italy and Spain, a narrow focus on contact tracing had missed widespread community transmission, leading to very rapid exponential growth thereafter. The slow linear growth over protracted periods in these 19 SSA countries may suggest effective containment, lower reproduction numbers, low health-seeking, and/ or a low ceiling on capacity for testing suspected cases. Limited testing capacity is a plausible explanation for the variable growth rates seen in some other countries. However, the exceptionally rapid progression from 1^st^ to 50^th^ case in those sub-Saharan African countries to have reached this threshold suggests that initial containment procedures may have not been as effective as in HIC. While most initial cases were confirmed in foreign visitors, and returning diaspora from high risk countries, Africa was the last world region to be affected, and the number of high risk countries and the probability that any such entrant was infected would have increased by the beginning of March 2020. Many SSA countries were slow to introduce quarantine procedures and/ or ban entry from affected countries. Quarantine protocols may not have been as stringent as in HIC. More infected visitors may have passed through to seed infection undetected. When cases were identified, resources for contact tracing would have been more limited, and less informed by testing given very limited capacity in the region at that time [1]. It is probably safe to assume, whether formally confirmed or not, that all 13 SSA countries with 50 or more confirmed cases will have well-established and widespread community transmission. The rapidity of growth in cumulative cases in the nine SSA countries to have reached >=100 cases is particularly worrying. This resembles more closely the pattern seen in Italy and Spain, than that in other European countries with more attenuated subsequent growth curves.

Doubling times for selected European countries, and Japan, South Korea and the USA have been estimated across the course of the outbreaks in those countries, with data smoothed using a moving average filter, with the primary purpose of comparing the success of different and differently timed containment measures [14]. At the beginning of March 2020, doubling times in Italy, UK, USA and Australia were all around 3 to 4 days. In Japan and Korea doubling times had already lengthened to eight to 10 days reflecting the relative success of their early containment approaches. Subsequently, doubling times have lengthened in Italy as it approaches the zenith of its outbreak, while doubling times have shortened further in USA and Australia.

## CONCLUSION

One month into the outbreak, sub-Saharan African countries are at a tipping point. Tracking growth rates will be important, and doubling times are a serviceable indicator for this purpose. Given the ominous signs in many countries, now is the time to consider introducing more wide-ranging population-level regulations to control transmission. Increased development and donor assistance are essential to support these efforts and mitigate their consequences for the most disadvantaged. Careful planning is needed, considering needs for information and guidance, economic assistance, food supply, essential healthcare, and security. Political will is also needed in SSA countries to ensure disaster budgets are used efficiently and effectively, avoiding dependency mindset, which can impede early response. While the World Health Organization and Africa CDC have been very active in providing technical and material assistance in the region, there is still an urgent need for material and systems support to scale up testing efforts, to provide personal protective equipment, and to do whatever can be done, urgently, to upscale critical care capacity.

## Data Availability

The data used for these analyses was compiled from the World Health Organization Coronavirus disease (COVID-19) outbreak situation data dashboard, and can be accessed from the the #COVID_SSA data archive maintained by the NIHR Global Health Research Unit for Health System Strengthening in Sub-Saharan African Countries at King's College London (ASSET) at https://healthasset.org/daily-update-on-covid-19-in-sub-saharan-africa/#1585902823071-84797f00-a69e

## ACKNOWLEDGEMENTS

This research was funded by the National Institute for Health Research (NIHR) Global Health Research Unit on Health System Strengthening in Sub-Saharan Africa, King’s College London (GHRU 16/136/54) using UK aid from the UK Government to support global health research. The views expressed in this publication are those of the authors and not necessarily those of the NIHR or the UK Department of Health and Social Care.

